# Serological diagnostic kit of SARS-CoV-2 antibodies using CHO-expressed full-length SARS-CoV-2 S1 proteins

**DOI:** 10.1101/2020.03.26.20042184

**Authors:** Rongqing Zhao, Maohua Li, Hao Song, Jianxin Chen, Wenlin Ren, Yingmei Feng, Jinwen Song, Ya Peng, Bin Su, Xianghua Guo, Yanjun Wang, Jingong Chen, Jianli Li, Hunter Sun, Zhonghu Bai, WenJing Cao, Jin Zhu, Qinlu Zhang, Yufei Sun, Sean Sun, Xinkun Mao, Junchi Su, Hui Chen, Ailiang He, Ronghua Jin, Le Sun

## Abstract

WHO has declared COVID-19 a pandemic with more than 300,000 confirmed cases and more than 14,000 deaths. There is urgent need for accurate and rapid diagnostic kits. Here we report the development and validation of a COVID-19/SARS-CoV-2 S1 serology ELISA kit for the detection of total anti-virus antibody (IgG+IgM) titers in sera from either the general population or patients suspected to be infected. For indirect ELISA, CHO-expressed recombinant full length SARS-CoV-2-S1 protein with 6* His tag was used as the coating antigen to capture the SARS-CoV-2-S1 antibodies specifically. The specificity of the ELISA kit was determined to be 97.5%, as examined against total 412 normal human sera including 257 samples collected prior to the outbreak and 155 collected during the outbreak. The sensitivity of the ELISA kit was determined to be 97.5% by testing against 69 samples from hospitalized and/or recovered COVID-19 patients. The overall accuracy rate reached 97.3%. Most importantly, in one case study, the ELISA test kit was able to identify an infected person who had previously been quarantined for 14 days after coming into contact with a confirmed COVID-19 patient, and discharged after testing negative twice by nucleic acid test. With the assays developed here, we can screen millions of medical staffs in the hospitals and people in residential complex, schools, public transportations, and business parks in the epidemic centers of the outbreaks to fish out the “innocent viral spreaders”, and help to stop the further spreading of the virus.

## Introduction

As of March 24, there were 370,416 confirmed cases of COVID-19 with 16,324 deaths in the world^1^. Infections among healthcare providers were even more alarming, with 4826 Italian doctors and nurses reported to be infected over such a short period due to the lack of appropriate medical protection gear and quick screening of SARS-CoV-2 infections^2,3^. Making the issue even worse, the virus can be widely transmitted by asymptomatic viral-carriers to people in close contact^4^, with some patients reportedly becoming sick once again after their initial recovery and yielding a positive NAT test^5^. There is an urgent need to develop rapid, fast and simple screening tools to find “moving viral carriers” and quarantine them.

Nucleic acid tests (NAT), the most widely used diagnostic assay for COVID-19 in the world, only have a 40% accuracy rate, leading to many patients who test negative on NAT to nonetheless go on to suffer severe complications, including possible mortality, due to undetected SARS-CoV-2 infections^6,7^.

SARS-CoV-2 is one of the seven human coronaviruses known to infect human. Out of the seven members, SARS, MERS and SARS-CoV-2 have mortality rates between 9-14%, while the other four members, HCoV-OC43, HCoV-NL63, HCoV-229E and HCoV-HKU1, only cause mild flu syndrome and have been around for many years. SARS-CoV-2 contains nucleoprotein (N-protein), spike glycoprotein (S-protein), envelope protein (E-protein) and membrane protein (M-protein)^8,9^. SARS-CoV-2’s N protein shared over 90% homology with other members of the coronavirus family. Cross-reactivity among N proteins of human coronaviruses was reported by Yu’s group^10^. The S-protein can be divided into two parts: S1 and S2 proteins. The S1 protein attaches the virion to the cell membrane by interacting with a host receptor (human ACE2), initiating the infection^11^. SARS-CoV-2’s S1 protein has 685 amino acids with seven potential glycosylation sites^12^. No evidence of strong cross-reactivity was observed with many neutralizing SARS or MERS monoclonal antibodies (personal communications), suggesting it has very unique antigenicity.

Several diagnostic kits for measuring SARS-CoV-2 IgM and IgG have been approved by Chinese FDA with the restriction that they may only be used as companion tests for NAT, and not to be used for general screening of SARS-CoV2 infection due to lacking the required specificity and sensitivity. Possible cause may be the poor quality of the detecting antigens used. Three different types of antigens were reported to be used: 1) the recombinant N protein from SARS-CoV-2, which is highly conserved among all 7 members of coronaviruses and led to poor specificity in tests of general population, 2) CHO-expressed S1 protein from SARS-CoV-2, which has very different antigenicity from its counterpart in SARS-CoV-2, or 3) the receptor binding domain (RBD) of SARS-CoV-2 S1, which is about 200 amino acids long with only one glycosylation, compared to the full length S1 which has 7 glycosylation sites. These latter two can result in poor sensitivities. Misdiagnose of HCoV-OC43, HCoV-NL63, HCoV-229E and HCoV-HKU1infections as SARS-CoV-2 could send thousands and thousands of people to already over-loaded hospitals and increase the risk of real infection by SARS-CoV-2 during the unnecessary hospital visit. On the other hand, missed detections of SARS-CoV-2 infections can deny patients the opportunity to receive early preventative care before the disease progresses into acute respiratory syndrome, which has an over 60% mortality rate. Therefore, it is extremely important to develop serological tests using the right detecting antigen: fully glycosylated, full length SARS-CoV-2 S1 recombinant protein(s).

The full length SARS-CoV-2 S1 protein has previously been difficult to express at a commercially viable level (personal communications), but using our patented technology, we have improved the expression level by close to a hundred fold (∼80mg/L) using either CHO or 293F mammalian cells. Using the CHO-expressed SARS-CoV-2 S1 protein as the detecting antigens, we have developed a very sensitive and highly specific diagnostic assay for screening the health care staff at the hospitals to reduce the in-hospital infections, and checking the in-coming visitors from the epidemic areas, and the work forces coming back to work, and the general populations for SARS-CoV-2 viral infection.

## Materials and methods

### Regents and supplies

Freund’s complete adjuvant (CFA), Freund’s incomplete adjuvant (IFA), Polyethylene glycol 4000 (PEG4000), DMSO, TMB substrates were purchased from SIGMA, USA. High-binding 96-well ELISA plates were purchased from Corning, USA. L-glutamine, antibiotics. ELISA buffers and solutions were prepared using analytic reagent-grade chemicals unless specified otherwise. Goat anti-human IgG (H+L) peroxidase conjugate was sourced from Jackson Immunoresearch, USA. Cynomolgus monkeys were hosted at Xierxing Biotech., Beijing, China. Mouse anti-His 6X mAb 6E2 was provided by AbMax. HEK 293F cells and CHO cells and culture media were provided by Zhenge Biotech., Shanghai, China. SDS-PAGE precast gels were purchased from GenScript, China.

### Protein expression and purification

The full length SRARS-CoV-2 S1 gene (GenBank: QIC53204.1) with C terminal were synthesized by GENEWIZ, China, and inserted into mammalian cell expression vectors with either 6* His tag or fused with human IgG Fc. The purified plasmid DNA was used to transfect mammalian CHO and human 293F cells by lipofection using liposome transfection kit (Invitrogen, USA) following manufacturer’s instructions. The transfected mammalian cells were grown at 37°C and 5% CO_2_ for a few days prior to harvesting. The harvested cells were pelleted by centrifugation at 4,000 rpm for 10 minutes. The recombinant S1-His6X protein was by immobilized metal affinity chromatography. The recombinant S1-Fc protein was purified by Protein A chromatography. Protein concentration was determined by OD absorbance at 280nm. The purity and identity of the purified recombinant SRAR-CoV-2 S1 proteins were ascertained by SDS-PAGE, Coomassie brilliant blue staining and ELISA with anti-6* His mAb. Briefly, the samples were loaded onto 12% gels, separated by SDS-PAGE, either stained with Coomassie brilliant blue staining for purity.

### Generation of monkey polyclonal antibodies

10 months-old Cynomolgus monkeys were first immunized with CHO-expressed S1-Fc fusion protein in Complete Freund’s Adjuvant and boosted in Incomplete Freund’s Adjuvant. Two to four weeks after the first immunization, bleeds were tested for titers by indirect Enzyme-linked Immunoassay (ELISA).

### Serum samples for assay

The protocols were approved by the institutional ethical committee of Beijing You An Hospital, Capital Medical University. Strong negative plasma samples and negative ones were obtained from human subjects that were collected prior to and during the COVID-19 outbreak respectively. Plasma samples were also obtained from hospitalized and/or recovered patients confirmed SARS-CoV-2 virus infection. Informed consent was obtained from all the human subjects who participated in the study after the nature and possible consequences of this study had been fully explained and the protocols were approved by the institutional ethical committee. The serum samples were inactivated at 56 °C for 30 min and stored at −20 °C until used.

### SARS-CoV-2 virus serology ELISA kit

Briefly, known amount of recombinant full length SARS-CoV-2 S1-His was diluted in PBS (10 mM, pH 7.4) and 100µL of the solution was added to each well of 96-well high binding ELISA plates (8 wells/strip, Corning, USA) and incubated overnight at 2-8°C. The wells were emptied and washed twice with PBS and unsaturated sites were blocked with 3% BSA in PBS by incubating for 1 hour at room temperature. Coated plates were air-dried and sealed in plastic bags and stored at 2-8°C until used.

Anti-SARS-CoV-2 S1-Fc monkey pAbs or human plasma sample were first diluted in negative human sera (pooled serum from 8 human subjects prior to the outbreak of COVID-19). For ELISA, each serum sample was tested in duplicates and 46 samples can be accommodated on one plate. Prior to test, human samples or the standards were diluted 1:20 in sample dilution buffer, such as 20% Calf-serum (CS) in PBS. 100µL of appropriately diluted sample was added to each well of the S1-His-coated plates and incubated for 1 hour at 37°C with constant shaking. The wells were emptied and washed twice with PBS containing 0.1% Tween 20 (PBST). 100 µL of appropriately diluted goat anti-human IgG (H+L) peroxidase conjugate in 20%CS in PBS was added to the respective wells and incubated for another 1 hour at 37°C with constant shaking. The wells were emptied and washed five times with PBST before addition of TMB substrate solutions. The chromogenic development was stopped using 0.1M H_2_SO_4_ after 15 minutes of incubation in the dark. Optical density (OD) was measured at 450nm wave length in a microplate spectrophotometer (Thermo Scientific, Multiskan MK3).

Calculate the mean value (AVG1) of Negative control, and times the lot-specific converting factor (CF) as the negative cut-off point (N-Cut); calculate the mean value (AVG2) of Weak positive control, use AVG2 as the positive cut-off point (P-Cut). If the absorbance value of the sample is greater than or equal to the positive cut-off point (P-Cut), the result of the sample is positive, indicating that the sample has detected antibodies that recognize the SARS-CoV-2; if the absorbance value of the sample is less than the negative cut-off point (N-Cut), the result of the sample is negative, it means that no antibody that recognizes the SARS-CoV-2 is detected in the sample; if the absorbance value of the sample is less than the positive critical point value (P-Cut) and greater than or equal to the negative critical point value (N-Cut), the result of the sample falls into a grey area and needs further experimental confirmation.

## Results

### Construction, expression and purification of recombinant SARS-CoV2 S1 proteins

The Spike protein S1 plays a key role in virus binding and entering host cells via human ACE2. It has 685 amino acids with 7 potential glycosylation sites, and its heavy glycosylation made it with very distinguishable antigenicity from its close family members SARS and MERS, demonstrated by the no significant cross reactivity was observed with existing neutralizing mAbs to SARS or MERS. The DNA sequence corresponding to the full length SARS-CoV-2 S1 protein was chemically synthesized and inserted into two different mammalian cell expression vectors with either 6XHis tag or human IgG Fc region to produce two versions of recombinant SARS-CoV-2 proteins, S1-His and S1-Fc (Fig. 1A).

**Figure 1.**
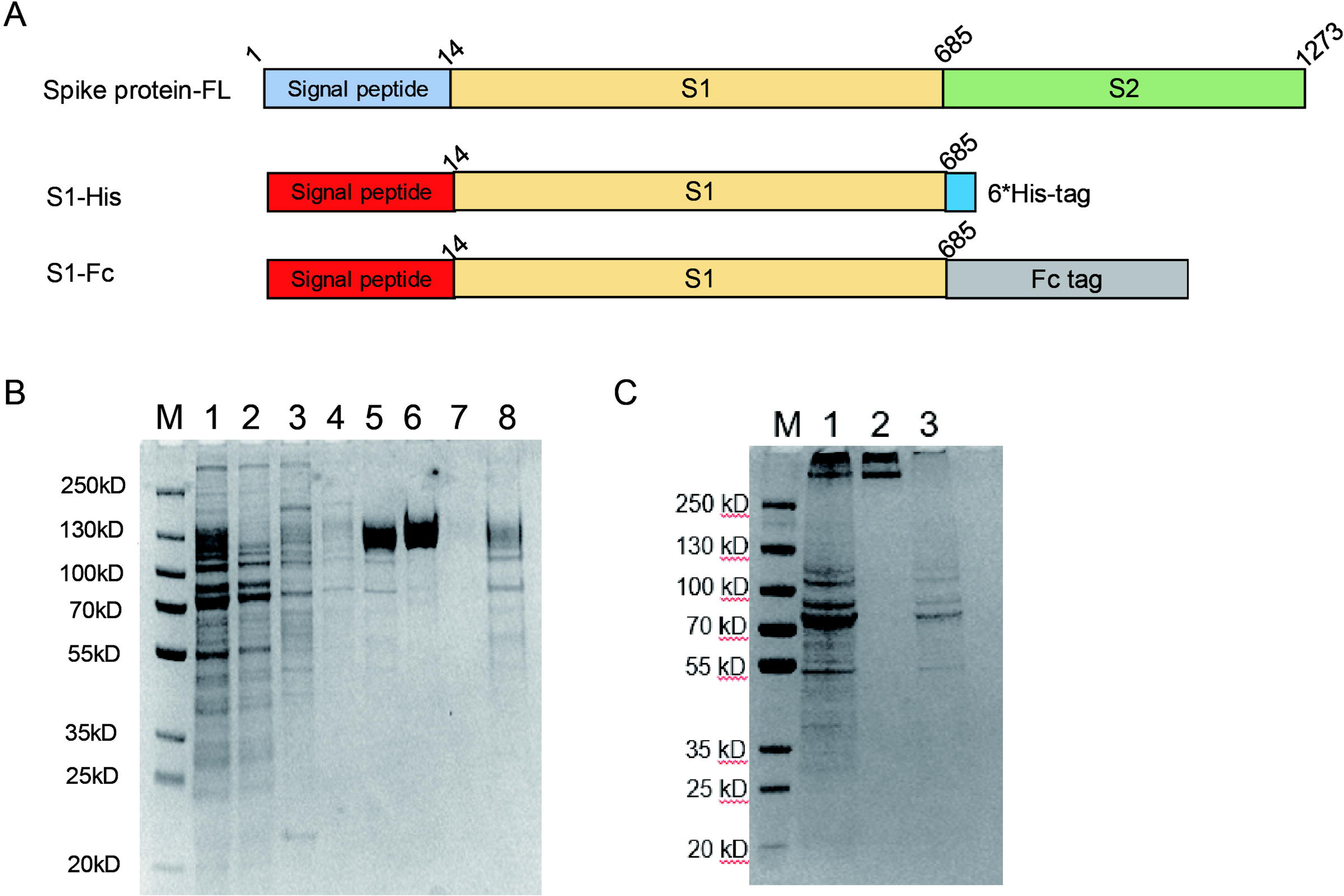
Construction and expression of the SARS-CoV-2 S1 protein. A) Domain structures of SARS-CoV-2 Spike 1 proteins, including full length spike protein S, S1 protein with 6* HIS and Fc TAG: the signal peptide was colored with blue or red, S1 domain was coloured with yellow, S2 domain was coloured with green, His tag was coloured with light-blue and Fc tag was coloured with grey. B) SDS-PAGE of S1-His expression and purification: Lane M referred to the MW markers (kDa), lane 1 referred to the culture supernatant, lane 2 referred to the flow-through, lane 3 referred to the 1st wash with buffer 1, lane 4 referred to the 2nd wash with buffer 2, lanes 5∼8 referred to three different fractions eluted with buffers containing 50 mM MES, 250 mM Imidazole, 150 mM NaCl pH 7.4. C) SDS-PAGE of S1-Fc expression and purification: Lane M referred to the MW markers (kDa), lane 1 referred to the culture supernatant, lanes 2 referred to eluted fractions, lane 3 referred to the flow-through.

Culture supernatants were purified using either Ni column or Protein A column. As shown in Fig.1B, a defused band was observed around 120 kD in elution (Lane 5 and 6), which is much larger than the expected size of S1-His, suggesting heavy glycosylation took place. Two sharp bands at 500 kD or higher were detected in the elution of Protein A column (Fig. 1C), representing dimer and oligomer of S1-Fc.

Multiple batches of expressions and purifications of both S1-His and S1-Fc recombinant proteins using two mammalian cell systems have been carried out. The culture supernatants and cell lysates were collected at different times and the expression levels were examined by ELISA using mouse anti 6* His tag mAb 6E2. Using our patented technology, the transient expression levels of S1-His and S1-Fc in either CHO cells or 293F cells reached 30-72mg/L (Table S1). Stable cell lines just established recently.

### Characterization of the purified recombinant SARS-CoV-2 S1 proteins

To verify the true identity, the purified recombinant S1-His protein was coated on to 96-well plate and examined with the plasma samples collected from recovered COVID-19 patients. As shown in Fig. 2A, all six plasma samples reacted strongly with the purified recombinant SARS-CoV-2 S1 protein, indicating not only the right sequence but also the correct conformation.

**Figure 2.**
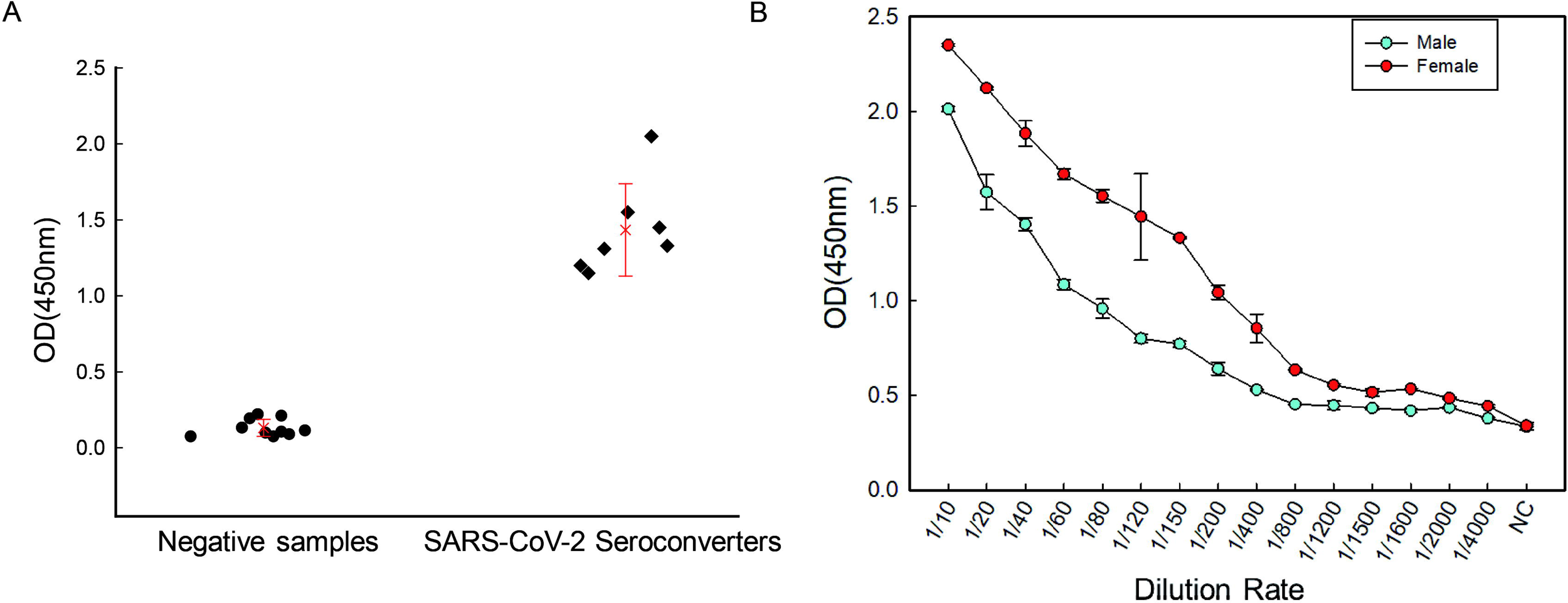
Identification of recombinant S1 protein. A) Recombinant S1-HIS protein was coated on the plate and characterized by plasma samples. The negative samples represented plasmas from non-infected populations and SARS-CoV-2 seroconverters indicated plasmas from recovered COVID-19 patients. B) Monkey sera after immunization with S1-Fc. Each well was coated with 100 µl of S1-His6X at 1 µg/mL, probed with different concentrations of monkey sera prepared in 20% CS-PBS. After washes, the immune complexes were detected with Goat anti-Human IgG (H + L) peroxidase conjugate (1: 20,000 in 20% CS-PBS). TMB substrate solution was added and OD measured at 450 nm wave length in a microplate spectrophotometer.

To generate the positive controls for the COVID-19 serological assays, two monkeys were immunized with recombinant S1-Fc with the help of adjuvant. As shown in Fig. 2B, on day 16, both monkeys developed very strong immune reactivity against S1-His, with titers at 1:200 for the male monkey and 1:800 for the female one. The two sera were mixed and used to spike the human normal sera for preparation of the positive controls.

### Development of Serological assays for SARS-CoV-2 antibodies

To evaluate the effect of S1-His protein coating concentrations for capturing anti-SARS-CoV-2 antibodies in testing samples, each well of 96-well EIA plate was coated with 100 µL of S1-His protein at eight different concentrations (0.1, 0.2, 0.4, 0.5, 0.8, 1.0, 1.2 and 1.5 µg/mL) in 10mM PBS pH 7.4 at 2-8°C overnight.

The solid phase-bound S1-His protein was probed using appropriately diluted mouse anti-S1-His mAb 6E2 (0, 2, 5, 20 µg/mL). As shown in Table S2, at all eight different coating concentrations of S1-His, the OD values showed dose dependency, while the maximal OD values increased with increasing coating concentrations of S1-His protein. Since the background OD values did not change significantly, 1.5µg/mL concentration of S1-His protein was considered optimal for coating of ELISA plate for kit manufacture to ensure the highest sensitivity.

There is a need to dilute testing human sera since matrix components, especially the host antibodies, can contribute to high assay background if undiluted. A set of 8 strong negative plasma samples (collected prior to the outbreak of COVID-19) were tested at five different dilutions to experimentally determine the assay’s optimal dilution.

As shown in Table S3, if the dilution is not high enough, such as 1:5 or 1:10, the background is too high. At 1:20 or higher dilutions, the background was acceptable. Base on suggestions from the clinicians, 1:20 dilution is more practical and determined to be the dilution rate for future use.

To balance between preservation of the detection of weak affinity SARS-CoV-2 antibodies and reduction of background, we have tried different washing, sample dilution and enzyme dilution buffers.

Addition of detergent for sure will reduce the non-specific binding of antibodies binding to the plates, but too much of it will also remove some of the blocking of the plate and give more opportunity for non-specific binding. As you can see from Fig. 3A, with detergent Tween-20 in the washing buffers, the background was significantly reduced.

**Figure 3.**
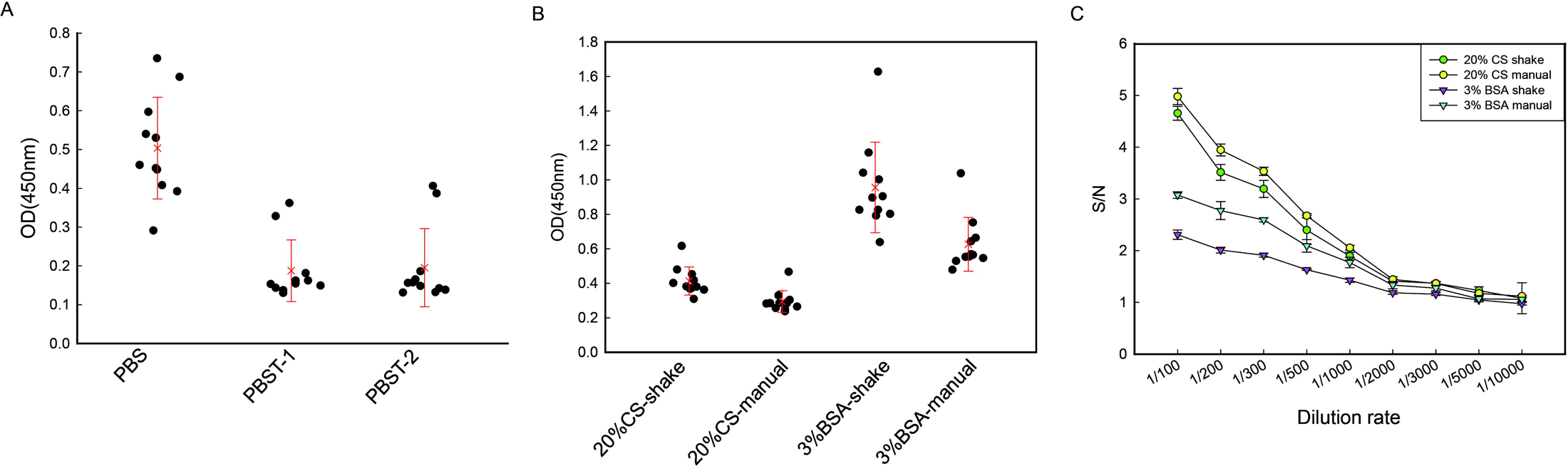
Kit optimization for the serological assay. A) Different washing buffers including PBS and PBST were tested. PBS (0.01M phosphate-salt buffer, pH 7.4), PBST-1 (PBS-0.05% tween-20), and PBST-2 (PBS-0.1% tween-20). B) ∼C) Optimization of sample dilution buffer and enzyme dilution buffer. Different buffers were chosen for optimization of sample and enzyme dilution buffers by negative samples (B) and positive controls (C). 20%CS referred to 20% (v/v) calf serum in 0.01M phosphate-salt buffer (pH 7.4), 3% BSA referred to 3% (3g/100ml) BSA in 0.01M phosphate-salt buffer (pH 7.4), shake referred to 96-well plate with constant shaking at 200rpm, and manual mean 96-well plate with manual tapings every 10 minutes.

We also examined the influences of the sample dilution buffers, enzyme dilution buffers and assay conditions on the performance of the tests. Human plasma or serum contains extremely high level of antibodies which will non-specifically bind to the wells and could potentially increase the background significantly. One approach is to use non-human sera, such as calf-serum (CS), to compete for the non-specific binding. In this study, we tested both 3% BSA-PBS and 20% CS-PBS as the sample dilution buffers against 8 normal human plasma samples, the human negative control, the strong human negative control and human sera spiked with various amounts of sera from SARS-CoV-2 S1-Fc immunized monkeys. In the same experiment, we also tried the incubation with or without the constant shaking. As shown in Fig. 3B & 3C, 20%CS-PBS significantly reduced the background, comparing to 3% BSA-PBS, with much better signal-to-noise (S/N) ratio. Although taping the plates every 10 minutes produced similar results as constant shaking, it is still highly recommended to use temperature controlled microplate shaker.

Based on above data, the best manufacturing and key testing parameters for the SARS-CoV-2 serological ELISA kit were selected as 1) 1.5μg/mL SARS-CoV2 S1-His for plate coating, 2) 1:20 dilution of human sera using 20% CS-PBS as sample and enzyme diluent, 3) incubation with constant rotation using a temperature controlled micro-plate shaker.

### Reproducibility: Intra-assay and inter-assay precisions

Several batches of the SARS-CoV2 serology ELISA kits were manufactured at three different locations, and were tested using positive monkey sera at different dilutions in human sera for assessing the reproducibility of manufacturing and assay precisions.

Summarized in Table S4, all three batch’s Intra-CVs were in the range of 6.39-12.05%, which is within the acceptable criterion of less than or equal to 15%. The intra-assay imprecision of samples (CV) was around 10.38%.

### Specificity of the serological ELISA assay

The diagnostic specificity of the kit was demonstrated by testing 412 human samples including 257 samples collected prior to (strong negatives) and 155 samples collected during (negatives) the outbreak of COVID-19.

As shown in Table 1, for the strong negatives, obtained from different sources including 48 samples from Rabies vaccinated patients, showed very similar specificities between 95.6-100%. In the group of Commercial #2, they were from 50 Blacks, 30 Whites, 24 Asian females and 20 Asian males, and no significant difference in background was observed between different races or genders. The specificity for strong negative was determined at 96.9%.

**Table 1.** Specificity and sensitivity assay against strong negative samples

| Groups | Sources | SARS-CoV-2 Ab | SARS-CoV-2 Ab | Sub | Specificity |
| --- | --- | --- | --- | --- | --- |
|  |  | Negative | Positive | Total |  |
| Strong<br>Negative | Rabies vaccinated | 47 | 1 | 48 | 97.9% |
|  | Commercial #1 | 20 | 0 | 20 | 100.0% |
|  | Commercial #2 | 119 | 5 | 124 | 96.0% |
|  | Hospital #3 | 43 | 2 | 45 | 95.6% |
|  | Clinical Lab | 20 | 0 | 20 | 100.0% |
|  | Total | 249 | 8 | 257 | 96.9% |
| Negative | Group #1 | 13 | 2 | 15 | 86.7% |
|  | Group #2 | 9 | 0 | 9 | 100.0% |
|  | Group #3 | 123 | 0 | 123 | 100.0% |
|  | Group #4 | 8 | 0 | 8 | 100.0% |
|  | Total | 153 | 2 | 155 | 98.7% |

Strong negative Samples were collected from prior to the outbreak of COVID-19 from different origins, negative samples were collected from during the outbreak of COVID from different cities of China. SARS-CoV-2 Ab Negative represented no SARS-CoV-2 antibodies were detected in the sample, SARS-CoV2 Ab Positive represented SARS-CoV-2 antibodies were detected in the sample.

For the negatives, group #1 was collected from Beijing, and groups #2-4 were collected from Zhejiang province, both areas have confirmed COVID-19 cases. In the initial test, 2 of the 15 samples from Beijing’s group were tested antibody-positive (Table 1). We performed the antigen competition assay using the rec. S1-His proteins at very high concentrations, and found that the signals could not be blocked, suggesting those two were false negatives. No positive was detected in the other three groups. The specificity was 98.7%. Combine the data from the strong negative samples, the overall specificity of the ELISA kit was 97.5% (402/412).

### Sensitivity of the serological ELISA assay

In collaboration with Chinese CDC, the ELISA kits were sent to several hospitals including two in Beijing and one in Wuhan to examine its sensitivity against the real clinic samples. Some of the data were presented in Fig. 4A-D. One study group encompass of 45 clinic samples from COVID-19 confirmed patients at different clinic stages at different ages with different genders. As shown in Fig. 4A, out of the 45 samples, 44 tested positive for SARS-CoV2 antibodies with a sensitivity of 97.7%. There were 21 samples (one on day-1, 3 on day-3, 7 each on days-4 and −5, 2 on day-6 and 1 on day-7) collected within one week of onset of COVID-19 diseases, all of them tested positive for SARS-CoV-2. So far, no significant difference in antibody levels observed between different genders or ages (Fig. 4B & 4C).

**Figure 4.**
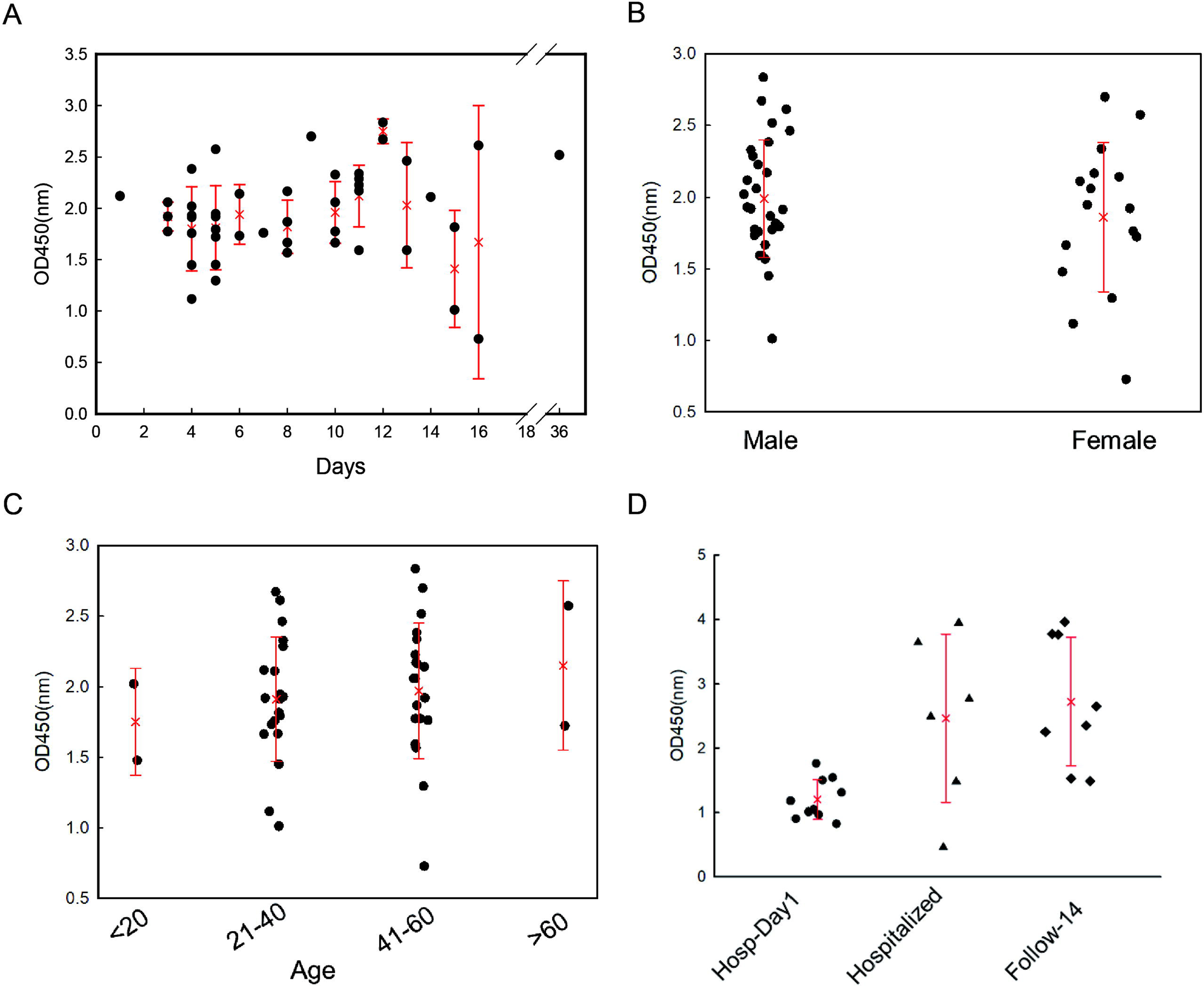
Sensitivity assay of the Kit. A) ∼C) 45 Plasma samples collected from either hospitalized or recovered COVID-19 patients were tested with the serological ELISA kit. A) days after onset of diseases, B) different genders, C) different age groups. D) 24 Plasma samples collected from either hospitalized or recovered COVID-19 patients, were tested with the serological ELISA kit. The samples were marked by collecting time: Hosp-Day 1 were samples collected one day after hospitalization, Hospitalized were samples collected anytime during the hospitalization, Follow-14 follow-up were samples collected on day 14 post-release from the hospital.

In another group of study, shown in Fig. 4D, 23 out of 24 clinic samples were tested positive for SARS-CoV-2 antibodies. We sort the samples by collecting time 1) one day after hospitalization (Hosp-Day 1), 2) anytime during the hospitalization (Hospitalized), 3) follow-up on day 14 post-release from the hospital (Follow-14). Clearly, the ones just arrived at the hospital had the lowest levels of SARS-CoV-2 antibodies. The antibody levels increased during the treatments and after been released from the hospitals. More works will be carried out to exam the levels of IgG and IgM of those positive samples respectively by simply changing the goat-anti-human IgG (H+L) secondary antibodies, which will detect both IgG and IgM, to goat-anti-human IgG Fc specific secondary antibodies and mouse anti-human µ Chain-specific mAb.

As summarized in Table 2, the overall sensitivity of the serological assay for SARS-CoV2 total antibodies was 97.1%.

**Table 2.**
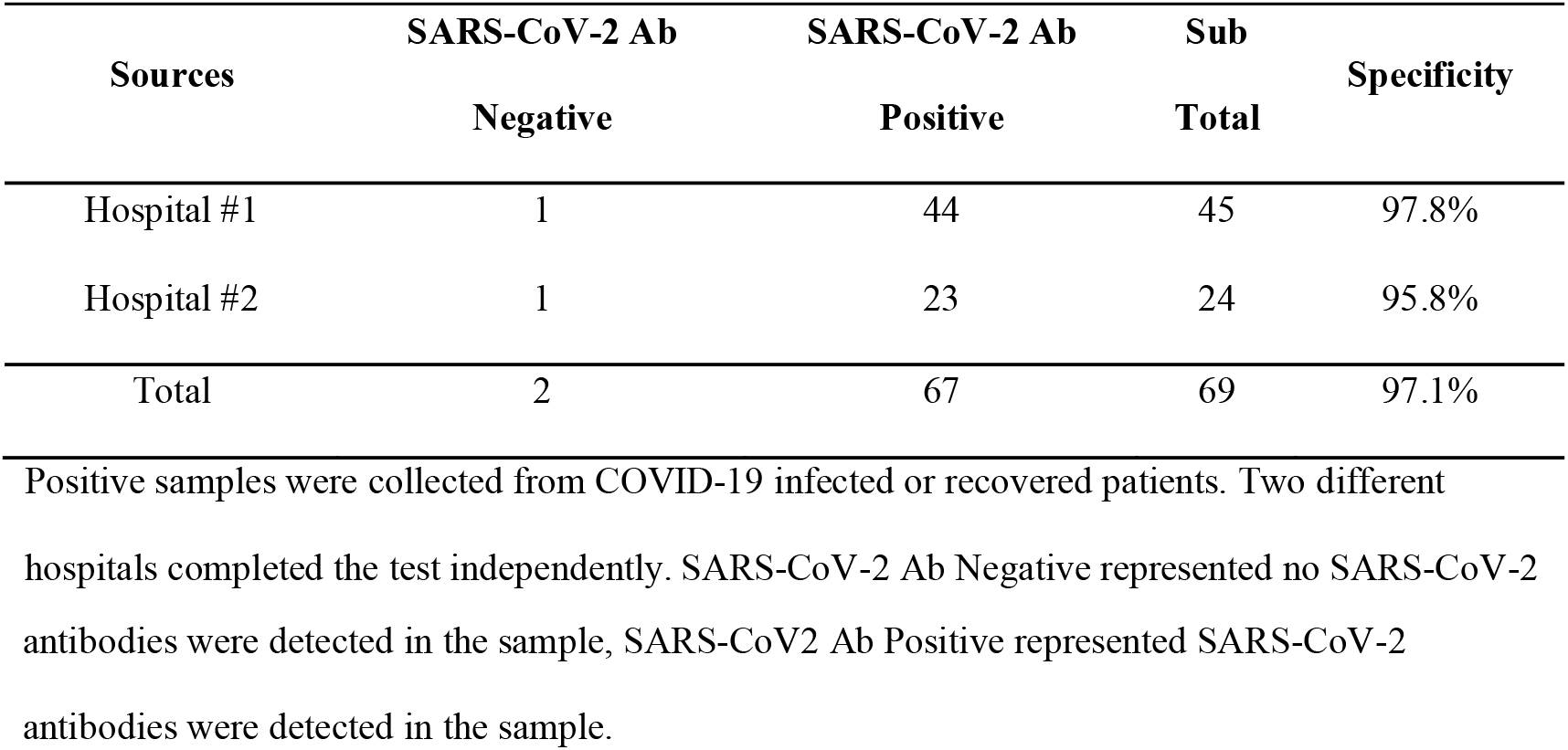
Sensitivity assay against positive samples

| Sources | SARS-CoV-2 Ab | SARS-CoV-2 Ab | Sub | Specificity |
| --- | --- | --- | --- | --- |
|  | Negative | Positive | Total |  |
| Hospital #1 | 1 | 44 | 45 | 97.8% |
| Hospital #2 | 1 | 23 | 24 | 95.8% |
| Total | 2 | 67 | 69 | 97.1% |

Positive samples were collected from COVID-19 infected or recovered patients. Two different hospitals completed the test independently. SARS-CoV-2 Ab Negative represented no SARS-CoV-2
antibodies were detected in the sample, SARS-CoV2 Ab Positive represented SARS-CoV-2 antibodies were detected in the sample.

Using the assay as screening tool for epidemiology study, in one case, there were five persons who were in close contact of confirmed COVID-19 patients and they had been quarantine for 14 days, showed no sign of sickness, tested negative twice by NAT and were released. One of them was tested positive for SARS-CoV-2 by this serological ELISA kit. This ELISA assay may offer a tool for the CDC teams who stayed in WuHan to search the clues for those new COVID-19 cases surfaced recently who had no clear connection with any confirmed COVID-19 patients.

## Discussion

The S1 protein binds to ACE2 protein on the surface of the human cells and plays critical role in virus infection. Our data showed that the S1 protein of SARS-CoV-2 virus is heavily glycosylated, evidenced by the purified S1-His protein which had an apparent molecular weight of 120 kD on SDS-PAGE gel, while its calculated molecular weight should be just around 70 kD. Glycosylation not only help the protein folding correctly, but also contribute greatly to protein’s affinity to its receptor. For example, the binding affinity of IgG1 to FcγRs on effector cell surfaces is highly dependent on the N-linked glycan at asparagine 297 (N297) in its CH2 domain^14,15^, with a loss of binding to the FcγRs observed in N297A point mutants^16,17^. Even the nature of the carbohydrate attached to N297 modulates the affinity of the FcγR interaction as well^18,19^.

In this study, full-length SARS-CoV-2 S1 proteins were expressed using both human 293F cells and Chinese hamster ovarian cells to ensure the recombinant proteins have the correct glycosylation profiles to resemble the native conformation on the surface of virus. Using our patented technology, we have successfully increased the expression levels of the full length recombinant SARS-CoV-2 S1 proteins up to 70mg/L. Using the CHO cell expressed full length SARS-CoV-2 S1-His protein as the capturing antigen, we have been able to develop a COVID-19 serological ELISA kit with high specificity (97.5%) and great sensitivity (97.1%). With an accuracy of 97.3%, the assay we developed here will be well suited for screening the health care staff to reduce in-hospital transfection of SARS-CoV-2 virus. Rapid Immunochromatographic Assay (RICA) was also developed using a double antigens Sandwich format. It is now in clinic study for its specificity and sensitivity. Once finished, we will have another tool to help patients to test at home so they either can receive early preventative care before the disease progresses into acute respiratory syndrome or not making un-necessary hospital visits for regular flu.

## Data Availability

The data used to support the findings of this study are available from the corresponding author upon request.

## Author contributions

RQZ, MHL, WLR, JXC, JGC, JLL, ALH contributed the development of recombinant proteins and assay development. WLR, YFS, ZHB, ZYS, QLZ, XKM, JCS, HC contributed in the pilot productions of assay kits. HS, YMF, XHG, JWS, YP, BS, YJW, WJC, JZ contributed the clinic studies. HS, SS contributed in market studies and technical writing.

## Acknowledgements

This work is supported by Research Grants from Beijing Science and Technology Commission, and Bill & Melinda Gates Foundation to Le Sun. This work was also supported by the National Natural Science Foundation of China (NSFC) (81702015), and the National Science and Technology Major Project (2018ZX10733403) to H.S. We thank Professor Wenjie Tan from National Institute for Viral Disease Control and Prevention, Chinese Center for Disease Control and Prevention for providing the sequences of SARS-CoV-2 S1 protien.

## References

1. “Reports of COVID-19 Pandemics, Confirmed cases and deaths in and outside of China.” www.163.com Mar. 24, 2020.

2. “Surging health care worker quarantines raise concerns as coronavirus spreads” www.khn.org Mar. 10, 2020.

3. “Thousands of medical staff infected by coronavirus in Italy” Gruppo Italiano per la Medicina Basata sulle Evidenze, Mar. 18, 2020.

4. “New cases of COVID-19 patients with no direct connections to confirmed patients surfaced at the clinics in Wuhan, a danger sign!” Health Daily, Mar. 17, 2020.

5. “Three members from the same family sick and NAT positive again. Expert cautioned maybe “false-cure”. Health Daily Mar. 21, 2020.

6. “Field Experts answered NAT will not be the only diagnose evidence” People’s Daily, Feb. 9, 2020.

7. “17year-old Korean boy died of COVID-19 but tested negative in 14 NAT” Korea Central Daily, Mar. 19, 2020.

8. Perlman, S. & Netland, J. Coronaviruses post-SARS: update on replication and pathogenesis. Nature reviews. Microbiology 7, 439–450, doi:10.1038/nrmicro2147 (2009).

9. Stadler, K. et al.. SARS--beginning to understand a new virus. Nature reviews. Microbiology 1, 209–218 (2003).

10. Yu, F., Le, M. Q., Inoue, S., Thai, H. T. C., Hasebe, F., Del Carmen, P. M. et al.. (2005). Evaluation of inapparent nosocomial severe acute respiratory syndrome coronavirus infection in Vietnam by use of highly specific recombinant truncated nucleocapsid protein- based enzyme-linked immunosorbent assay. Clin. Diagn.Lab.Immunol. 12, 848–854.

11. Stertz, S. et al.. The intracellular sites of early replication and budding of SARS- coronavirus. Virology 361, 304–315, (2007).

12. Wong, S. K., Li, W., Moore, M. J., Choe, H. & Farzan, M. A 193-amino acid fragment of the SARS coronavirus S protein efficiently binds angiotensin-converting enzyme 2. The Journal of biological chemistry 279, 3197–3201 (2004).

13. Wu, F., Zhao, S., Yu, B., Chen, Y.-M., Wang, W., Hu, Y., Song, Z.-G., Tao, Z.-W., Tian, J.-H., Pei, Y.-Y., Yuan, M.L., Zhang, Y.-L., Dai, F.-H., Liu, Y., Wang, Q.-M., Zheng, J.-J., Xu, L., Holmes, E.C. and Zhang, Y.-Z. A novel coronavirus associated with a respiratory disease in Wuhan of Hubei province, China. Unpublished. ACCESSION YP_009724390

14. Jefferis, R. & Lund, J. Interaction sites on human IgG-Fc for FcgammaR: current models. Immunology letters 82, 57–65 (2002).

15. Arnold, J.N., Wormald, M.R., Sim, R.B., Rudd, P.M. & Dwek, R.A. The impact of glycosylation on the biological function and structure of human immunoglobulins. Annual review of immunology 25, 21–50 (2007).

16. Shields, R.L. et al.. High resolution mapping of the binding site on human IgG1 for Fc gamma RI, Fc gamma RII, Fc gamma RIII, and FcRn and design of IgG1 variants with improved binding to the Fc gamma R. The Journal of biological chemistry 276, 6591–6604 (2001).

17. Tao, M.H. & Morrison, S.L. Studies of a glycosylated chimeric mouse-human IgG. Role of carbohydrate in the structure and effector functions mediated by the human IgG constant region. Journal of immunology 143, 2595–2601 (1989).

18. Kaneko, Y., Nimmerjahn, F. & Ravetch, J.V. Anti-inflammatory activity of immunoglobulin G resulting from Fc sialylation. Science 313, 670–673 (2006).

19. Shields, R.L. et al.. Lack of focus on human IgG1 N-linked oligosaccharide improves binding to human Fc gamma RIII and antibody-dependent cellular toxicity. The Journal of biological chemistry 277, 26733–26740 (2002).

